# Noncanonical EEG-BOLD coupling by default and in schizophrenia

**DOI:** 10.1101/2025.01.14.25320216

**Authors:** Michael S. Jacob, Brian J. Roach, Daniel H. Mathalon, Judith M. Ford

## Abstract

Neuroimaging methods rely on models of neurovascular coupling that assume hemodynamic responses evolve seconds after changes in neural activity. However, emerging evidence reveals noncanonical BOLD (blood oxygen level dependent) responses that are delayed under stress and aberrant in neuropsychiatric conditions. To investigate BOLD coupling to resting-state fluctuations in neural activity, we simultaneously recorded EEG and fMRI in people with schizophrenia and psychiatrically unaffected participants. We focus on alpha band power to examine voxelwise, time-lagged BOLD correlations. Principally, we find diversity in the temporal profile of alpha-BOLD coupling within regions of the default mode network (DMN). This includes early coupling (0-2 seconds BOLD lag) for more posterior regions, thalamus and brainstem. Anterior regions of the DMN show coupling at canonical lags (4-6 seconds), with greater lag scores associated with self-reported measures of stress and greater lag scores in participants with schizophrenia. Overall, noncanonical alpha-BOLD coupling is widespread across the DMN and other non-cortical regions, and is delayed in people with schizophrenia. These findings are consistent with a “hemo-neural” hypothesis, that blood flow and/or metabolism can regulate ongoing neural activity, and further, that the hemo-neural lag may be associated with subjective arousal or stress. Our work highlights the need for more studies of neurovascular coupling in psychiatric conditions.

## INTRODUCTION

Functional magnetic resonance imaging (fMRI) relies on models of neurovascular coupling to infer neural activity from fluctuations in blood oxygen levels. This model, known as the canonical hemodynamic response function (HRF), helps to delineate the spatiotemporal extent of tissue activation or deactivation in response to stimuli, tasks, and cognitive processing. According to this model, an increase in neural activity evokes cerebral blood flow to supply oxygen, glucose, and critical metabolites to active regions. Typically, the hemodynamic response evolves over the course of several seconds, peaking around 4-6 seconds in event related paradigms. However, this model makes assumptions that may not hold for all brain regions and particularly in the absence of an explicit task, such as during resting state studies (for reviews, see (1,2)). Therefore, the ongoing dynamics of neurovascular coupling may be more “noncanonical” than previously thought, although the extent of deviation from this model has not been fully explored.

The default mode network (DMN) is an exemplary resting-state network, generally defined by increased BOLD activity in the absence of an explicitly instructed task or stimulus (3–5). There is increasing evidence that regions of the default mode network demonstrate noncanonical hemodynamic coupling (6). Recent work, based on combined FDG-PET and BOLD-fMRI investigations, revealed “uncoupling” between BOLD deactivations in the DMN and unexpected increases in the cerebral metabolism of glucose. Paralleling the literature in non-human animals, intrinsically evoked (from spontaneous, ongoing activity) and extrinsically evoked (from an external stimulus) activity may yield different vascular and metabolic responses (7–10). This basic work extends evidence for noncanonical neurovascular coupling in the DMN, including carbon dioxide challenge paradigms (11), reduced cerebrovascular reactivity in aging (12), hypoxia (13,14) and under stress (15).

These empirical findings underscore the importance of metabolic factors in neurocognitive function and neuroimaging methods in which any singular experimental modality yields an incomplete picture of brain function. We have previously proposed an “active” role for metabolic factors in modulating ongoing neural activity, and that could drive intrinsically evoked BOLD activity in the absence of an explicitly cued task or stimulus (what we have referred to as *in*voked activity (16)). We extend the work of others who have linked intrinsic activity in the default mode network to the ongoing sense of “self-in-context” (17) to consider that BOLD activity could be uniquely invoked by arousal systems to prime neural systems for subsequent evoked responses. Our proposal is an extension of the hemo-neural hypothesis (18), that also inverts the evoked, hemodynamic recruitment model, in the sense that hemodynamic activity might evoke neural recruitment. These hypotheses may be of high relevance to neuropsychiatric conditions, where there is evidence of noncanonical hemodynamic responses. We have previously focused on schizophrenia, given evidence for abnormal hemodynamic responses (19,20) , impaired neuroenergetic coupling (21), and deficient EEG-BOLD coupling (22).

Therefore, work is needed to investigate noncanonical hemo-neural coupling in human participants. Our prior examination of resting alpha rhythms, from traditional hemodynamic modeling in psychiatrically unaffected participants, revealed minimal recruitment of the default mode network, instead revealing widespread inverse correlations with the visual and sensorimotor system (23). Our approach differed from prior simultaneous EEG-fMRI paradigms in that we examined resting rhythms relative to the aperiodic signal (24). Here, we re-analyze this EEG-fMRI data to examine the time course of EEG-BOLD coupling from a lagged-correlation analysis (25,26). We focus on the alpha rhythm and include participants with schizophrenia. The primary investigation is an exploratory, “model-free” analysis from lagged correlation between alpha fluctuations and whole-brain, voxel-wise BOLD dynamics. We hypothesized that alpha would be correlated with regions of the DMN, but in early temporal windows that don’t align with the traditional HRF. We further hypothesized delayed hemodynamic coupling in participants with schizophrenia. In general, we find evidence for noncanonical alpha-BOLD coupling within the DMN of all participants and that this coupling is delayed in schizophrenia. A subset of participants completed self-reported measures of stress, for whom delayed coupling was attributable to greater perceived stress levels, for both groups in the subgenual region of the ACC. These findings reveal a dynamic relationship between ongoing fluctuations in alpha power and the hemodynamics of the DMN than typically assumed. Further, they warrant re-examination of a “hemo-neural” hypothesis (18), whereby stress and arousal systems invoke hemodynamic activity to modulate ongoing neural activity and new neuroimaging methods that can accommodate these alternative models.

## METHODS

### Participants

The demographic and clinical data from schizophrenia (SZP; n=57) and unaffected participants (UAP; n=46) in this study has been previously reported (22,23). For full details of inclusion and exclusion criteria see (22). In brief, exclusion criteria were substance abuse in the past 3 months, any significant medical or neurological illness, or head injury resulting in loss of consciousness. In addition, UAP were excluded for any current or past psychiatric disorder based on the SCID-IV, any history of substance dependence (except nicotine), or having a first-degree relative with a psychotic disorder. We excluded SZP participants if they met criteria for substance dependence within the last year. All participants provided written informed consent based on procedures approved by the University of California at San Francisco and the San Francisco Veterans Affairs Medical Center Institutional Review Boards.

### EEG-fMRI acquisition

The details of our resting state, EEG-fMRI acquisition protocol and preprocessing pipeline have been previously reported in detail (27,28). We collected both structural and functional MRI data using a 3T Siemens Skyra scanner during an AC-PC aligned echo planar imaging (EPI) sequence. The fMRI resting-state scan duration was 6 minutes and pre-scan instructions requested participants to keep their eyes open and fixate on a white cross displayed on a black screen. An MR-compatible EEG cap (Brain Products) was used to record data from 32 scalp sites during MR-scanning. An additional electrode placed on the upper back to monitor ECG.

### ECG-EEG-fMRI Preprocessing

Structural and functional image preprocessing was conducted using Statistical Parametric Mapping 8 (SPM8; software) including motion correction and slice-time correction. We then implemented aCompCor (anatomic component based noise correction), a principal components-based approach for noise reduction (29). ECG and EEG data preprocessing first involved subtraction of the MR gradient artifact using Brain Vision Analyzer 2.0.4.368 software (BrainProducts) based on the methods of (30). The cardioballistic artifact removal was then performed in Brain Vision Analyzer 2 (BrainProducts, Gilching, Germany) based on the identification of R-peaks in the ECG that exhibited high temporal correlation (r > 0.5) with a heartbeat template and confirmed by visual inspection (31). Next, we utilized canonical correlation analysis (CCA) to remove electromyographic noise (32,33). Examples of our cardioballistic and EMG artifact correction procedures are available in (22). Lastly, data were re-referenced to an average reference and subject to independent components analysis (ICA) in EEGLAB. EEG data during artifactual TR intervals were removed based on FASTER criteria, spatial correlations with eyeblink and ballistocardiac artifact templates (34).

### Alpha Parameter Estimation

Our analytic pipeline is shown in Figure 1. Given the midline, posterior concentration of alpha power in both groups we focused our analysis on electrode Pz. Data were segmented into 2-second epochs aligned to the volume markers provided by the MR-scanner. Given that alpha power can be conflated with aperiodic activity, we sought to extract residual alpha power above the aperiodic backbone. Similar to our prior analysis (28) and following the methods of (24) we first performed a Fourier analysis using a 2-second Hanning window (0.5 Hz resolution) at each epoch. The aperiodic slope was fit within the range of 1-50 Hz and we examined residual alpha power within a band range of 8-12 Hz. Group differences in alpha power were determined from unpaired t-tests.

**Figure 1.**
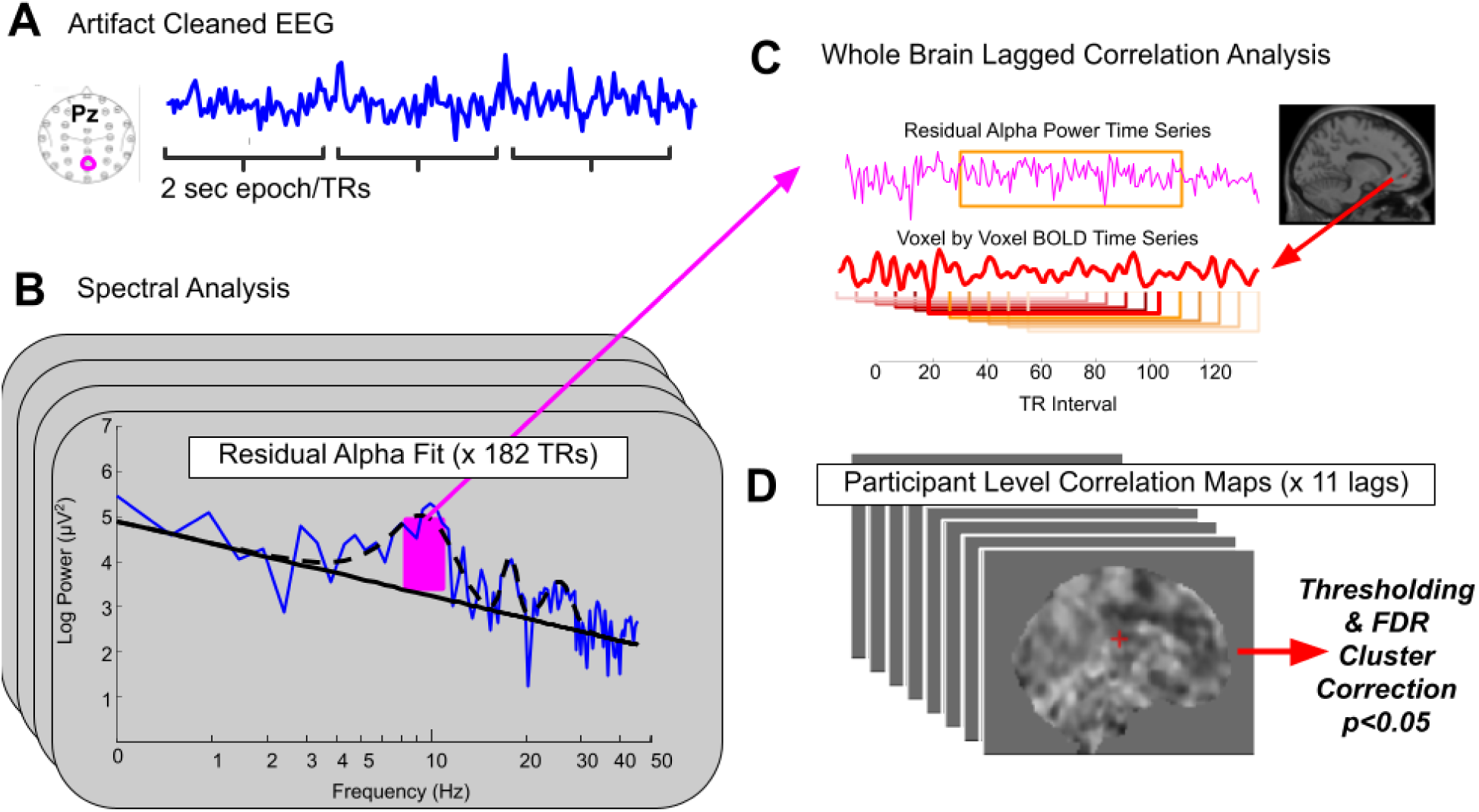
Analytic pipeline. (A) Artifact cleaned EEG, recorded simultaneously with fMRI, is epoched according to the 2 sec TR interval of the scanner sequence. (B) Spectral decomposition of the EEG (blue) is utilized to extract residual alpha power (magenta), rising above the aperiodic fit (solid black) for each TR epoch. (C) The timeseries of residual alpha power (magenta) is entered into a whole brain, lagged correlation with the timeseries of BOLD signal at each voxel (11 lags, -5 TR to + 5 TR) (D) Resulting correlation maps are thresholded and subject to FDR cluster-level correction over the 11 lags.

### Whole-Brain Lagged Correlation Analysis

Lagged correlation between alpha power and whole-brain, voxel-wise BOLD activity for each participant was conducted in the CONN toolbox (35). We lagged the residual alpha power from -10 seconds to +10 seconds, with 2 second spacing (corresponding to our TR) to yield 11 lagged correlation models. In effect, we use the different time lagged signals of alpha as a “seed” to identify the correlation coefficient between the EEG signal and all voxels in the brain. We set an initial cluster-finding threshold of p = 0.001 (two-tailed) and a spatial extent of 10 voxels for all models. For each lag value, we retained all clusters surviving this threshold, followed by FDR correction for multiple comparisons. To interrogate these maps further, we parcellated significant clusters according to the automated anatomical labeling atlas 3 AALv3 (36). Since this atlas does not include brainstem structures, we included an additional mask for voxels from this region, including the midbrain, pons and medulla. Only regions of size > 50 voxels after intersection with our significant clusters were retained for subsequent analysis.

### Accumulated Correlation Asymmetry

To assay the temporal asymmetry in alpha-BOLD coupling across regions and participants we calculated accumulated correlation asymmetry (ACA, (37)) for all significant brain regions. ACA “accumulates” (sums) cross-correlation coefficients on the right side (0s to 10s) and subtracts the sum of coefficients on the left side (-10s to 0). If the correlation peak is centered at zero lag, these coefficients should be equal and the difference should be 0. Under a canonical HRF, alpha-BOLD relationships should show positive ACAs, indicating that most of the correlated activity occurs when the BOLD signal lags alpha. ACA values were subject to a repeated measures anova, using a within participant factor of REGION (significant brain regions from the AALv3) and a between participant factor of GROUP (unaffected or schizophrenia). Regions with significant GROUP x REGION effects were further examined for correlation with clinical symptoms.

### Correlations with Stress and Symptoms

Given evidence for delayed hemodynamic coupling under stress, we also examined correlations between regional ACA scores and self-reported measures of stress (perceived stress scale, (38)) which was available from a subset of participants who had participated in a related neuroimaging study in our laboratory (15/46 unaffected and 22/57 schizophrenia participants). This subset of participants also reported their body mass index (BMI). To examine correlations between regional ACA scores and symptoms we utilized clinician ratings from the PANSS (Andreasen, 1982). Chlorpromazine equivalents were calculated using the minimum effective dose method (39,40) 5 participants in the SZP group were unable to confirm their dose at the time of the study and were excluded from those analyses.

## RESULTS

### Noncanonical Alpha-BOLD Coupling

At rest, SZP and UAP show no differences in residual alpha power (t=0.24, p=0.81; Figure 2). Across all participants and time-lags, we identify several highly significant clusters with alpha-BOLD correlations (FDR p<0.001; Figure 3A). In contrast with our prior findings using traditional HRF modeling, lagged-correlation analysis of alpha-BOLD coupling yields robust coupling in many of the canonical regions of the DMN. Several non-hypothesized regions also showed significant clusters, including the brainstem, cerebellum, thalamus, and caudate (Figure 3B). We find a diversity of alpha-BOLD temporal coupling profiles. This includes regions of the posterior DMN, which shows maximal correlation near 0-2 seconds lag. By contrast, regions of the anterior DMN are maximally coupled at BOLD lags of 6 seconds, closer to the latency of the canonical HRF. To interrogate these clusters further, and examine differences between groups, we parcellated them according the the AALv3, which identified 48 regions for further analysis.

**Figure 2.**
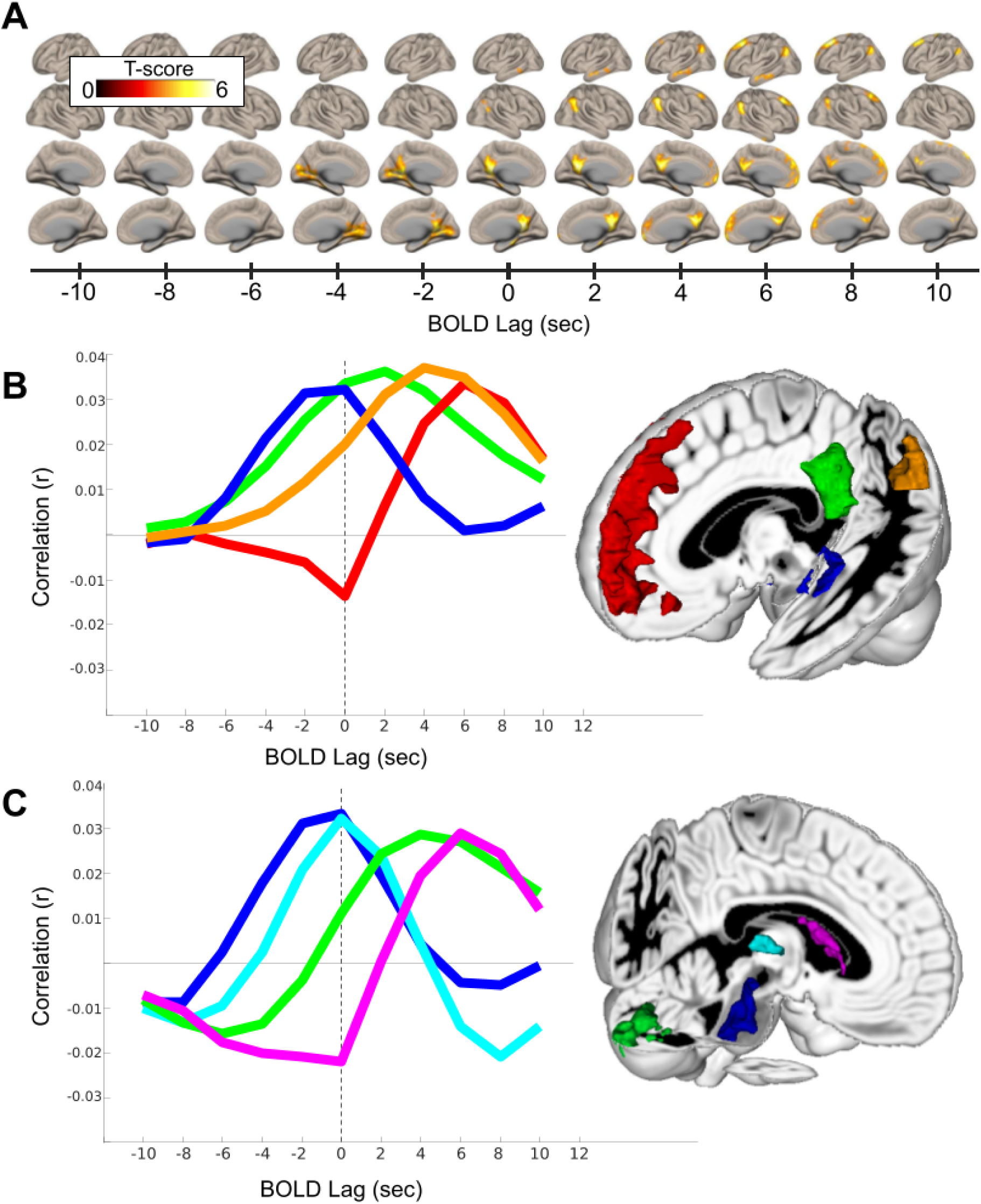
(A) T-scores from the average correlation maps across all participants, plotted on the surface of an inflated cortex. Each column represents the statistical strength of the correlation between residual alpha power and the BOLD signal for the lag value given by the abscissa. (B) Timecourse of alpha-BOLD coupling for 4 clusters overlapping with regions of the default mode network. Red: medial prefrontal regions, green: posterior cingulate/precuneus, orange: angular gyrus, blue: hippocampal gyrus. (C) Timecourse of alpha-BOLD coupling for 4 clusters from non-cortical regions. Purple: caudate, green: cerebellum, cyan: thalamus, blue: brainstem.

**Figure 3.**
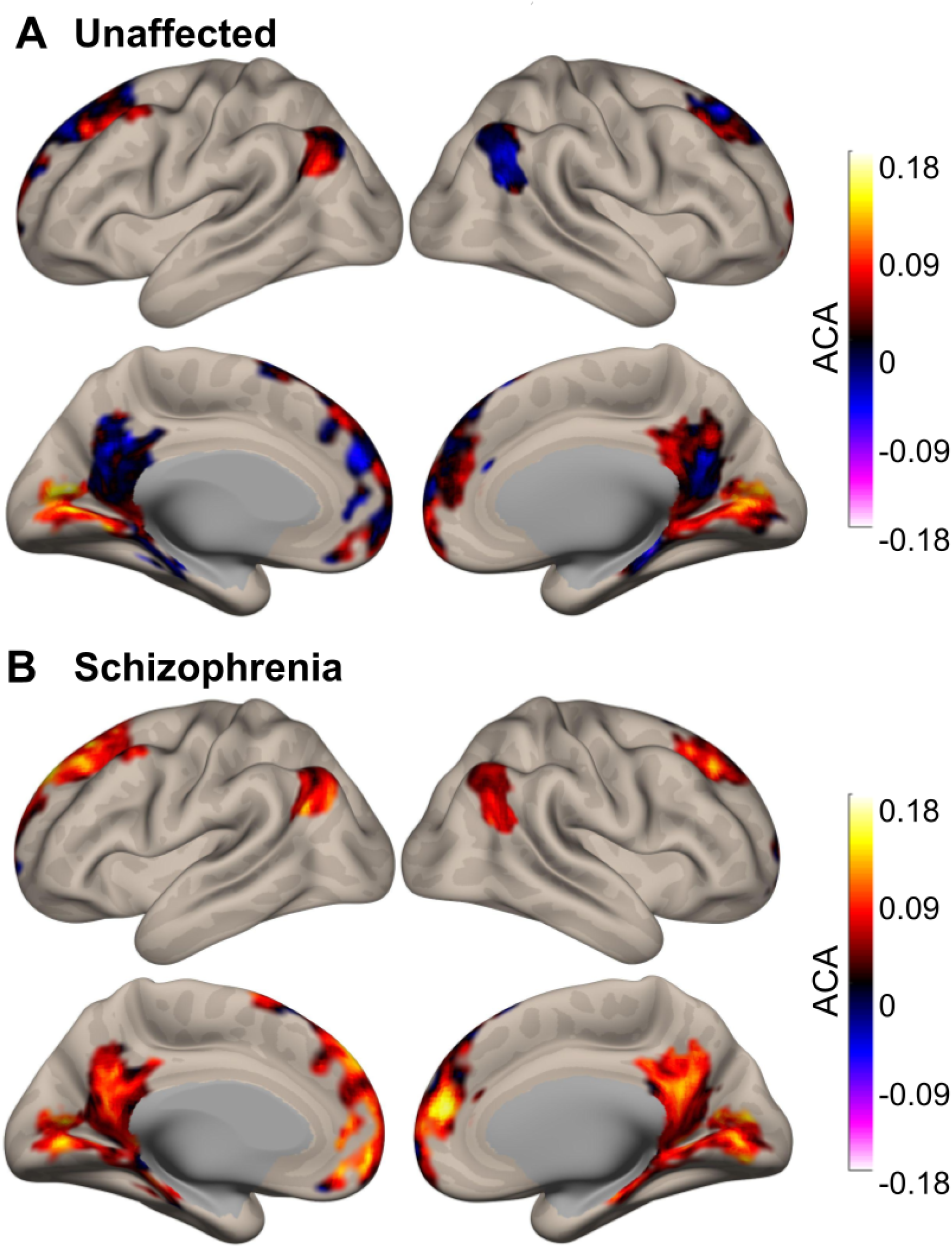
Maps of Accumulated Correlation Asymmetry (ACA) for unaffected participants (A) and schizophrenia participants (B).

### Alpha-BOLD Correlation Asymmetry in Schizophrenia

To assay the temporal profiles of coupling dynamics between UAP and SZP participants, we measured accumulated correlation asymmetry (ACA) for each region (see methods). ACA determines the magnitude of temporal precedence between signals, in our case, the degree to which the BOLD signal lags Alpha. The greater the value of ACA values, the greater the magnitude of BOLD correlation at positive lags (rightward shift). We measured ACA for all 48 brain regions, to compare coupling between UAPs and SZPs (Figure 4). We find a GROUP x REGION effect (F(47,4747)=1.82, p=0.0005) which is attributable to larger ACA values for SZPs in 12 regions (see Table 1). This indicates that alpha-BOLD coupling in SZP is right-shifted; BOLD signals lag ALPHA by greater magnitude.

**Figure 4.**
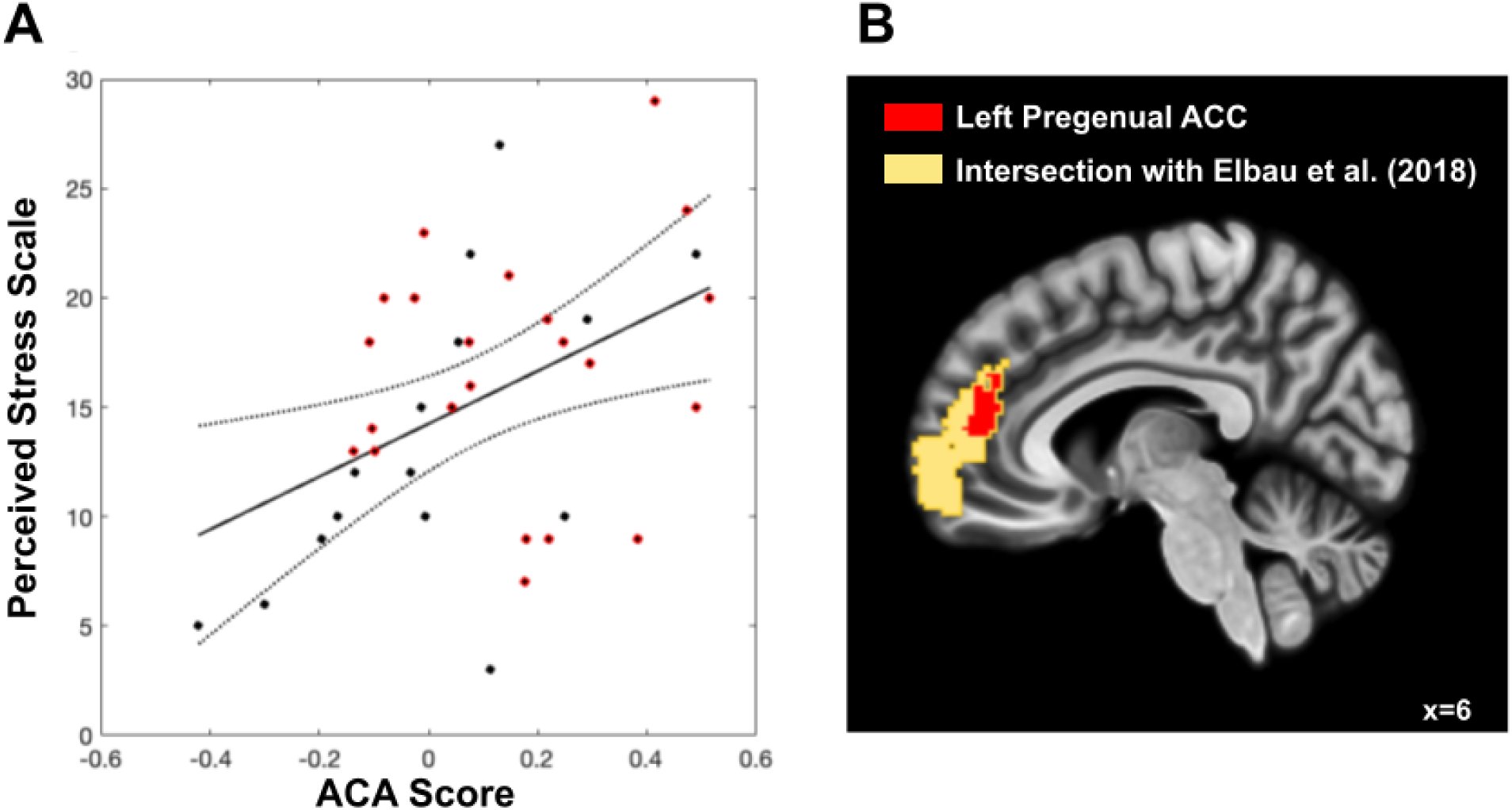
(A) Correlation between ACA score from the L Pregenual ACC cluster and total score from the perceived stress scale (r=0.43). Schizophrenia participants are labeled in red and unaffected participants are labeled in black. (B) Overlap between our L Pregenual ACC cluster medial prefrontal regions with delayed hemodynamic responses identified by Elbau et al. in their stress paradigm.

**Table 1.**
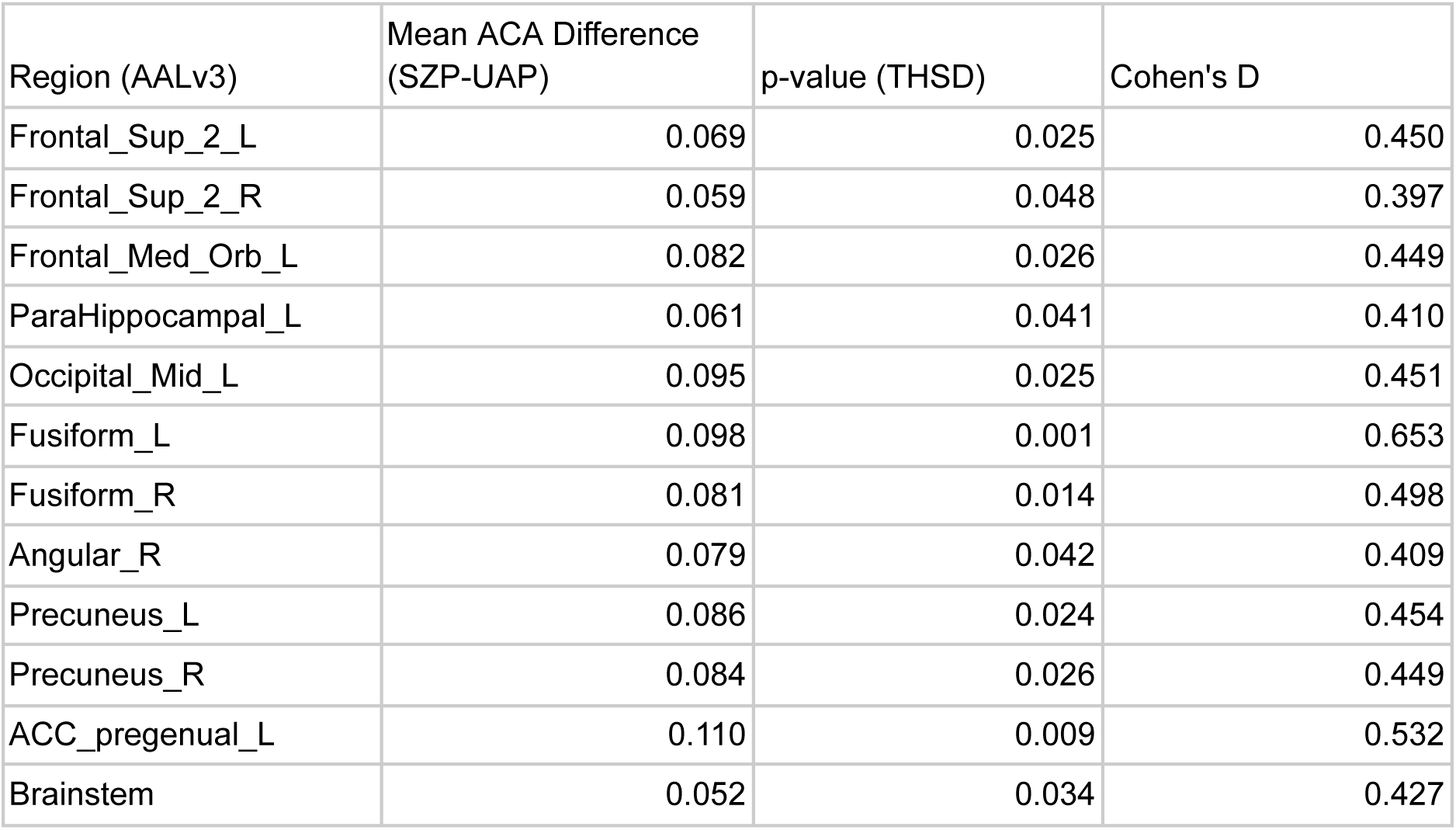
Regions with significant differences in ACA scores between UAP and SZP. Reported p-values reflect correction for multiple comparisons by Tukey’s Honestly Significant Differences (THSD). Effect sizes are given by Cohen’s D.

Given that the physiology underlying the variability in BOLD coupling is not understood, we applied additional tests to examine potential confounders, including age, gender, antipsychotic dose, body mass index (BMI) and heart rate. This interaction effect remained significant after controlling for the effect of age (F(47,4700)=1.76, p=0.001) and gender ( (F(47,4747)=1.82, p=0.0006), although post-hoc tests for Frontal_Sup_2_R, ParaHippocampal_L and Occipital_Mid_L were no longer significant after controlling for age. We found no significant correlations between ACA scores from the identified brain regions and chlorpromazine dose equivalents (all corrected p’s> 0.82), BMI (all corrected p’s> 0.19) and heart rate (all corrected p’s> 0.33).

### Relationship to Stress and Symptoms

Given existing evidence for delayed hemodynamic responses under stress, we further hypothesized that greater ACA values might be associated with levels of perceived stress. Although participants were not explicitly stressed by our experimental paradigm, a subset of participants completed self-report measures of stress from the perceived stress scale during a separate study conducted within our research group. We restricted our analysis to those regions that also show stress specificity from Elbau et al. (15), including the right hippocampus, bilateral anterior cingulate cortex, and medial orbitofrontal cortex. We find that self-reported stress is associated with ACA in the pregenual region of the anterior cingulate cortex across all participants (left: r=0.43, p=0.008; right: r=0.34, p=0.040). There was no difference in perceived stress between UAP and SZP (t=-1.24, p=0.22), nor was there a difference between groups in the direction or magnitude of the association between perceived-stress and ACC ACA (left: p=0.10; right: p=0.15). We found no correlations between ACA scores from any significant brain regions and positive or negative symptoms (all corrected p’s> 0.74).

## DISCUSSION

We have employed simultaneous, resting-state EEG-fMRI to examine coupling between alpha power and the BOLD signal. Lagged correlation analysis between these signals yields pronounced temporal variability, particularly within the canonical regions of the DMN. For posterior regions, including the posterior cingulate and hippocampal gyrus, alpha lags, or is nearly coincident with BOLD activity, inverting the standard assumptions of a hemodynamic delay. Anterior regions, including those of the medial prefrontal cortex, show canonical BOLD lags of 4-6 seconds. Furthermore, we found that BOLD lags are more pronounced in SZP, particularly the pgACC, fusiform gyrus and bilateral, superior frontal regions. Lastly, we found that the magnitude of the BOLD lag in the pgACC was associated with subjective reports of perceived stress.

Our findings build upon a growing literature supporting regional variability in EEG-BOLD coupling, including that in certain cases, BOLD activity may precede and predict EEG, rather than BOLD being evoked by EEG. Thus, depending on the region, the context, or the participant, the BOLD signal may come earlier or later than expected, relative to ongoing neural fluctuations. We substantiate prior findings of hemodynamic delays in stress and schizophrenia, by directly linking those delays to neural activity, through simultaneous EEG-fMRI. In the sections that follow, we discuss potential mechanisms and functional significance of these findings, as well as the implications for neuroimaging methods, particularly for neuropsychiatric populations.

### BOLD Variability and the Default Mode Network

Although variability in the HRF has been studied extensively in task-based paradigms, new computational methods for resting-state studies in addition to multimodal imaging methods have improved our ability to detect the physiology of intrinsic variability in BOLD signals (1,2). Prior studies have also utilized simultaneous EEG-BOLD and lagged correlation to examine coupling across the whole brain (26,41). Feige et al. report wide ranging temporal dynamics in alpha-BOLD coupling, albeit at a 5 sec TR, so the temporal resolution is difficult to parse.

Nonetheless, they find positive alpha correlation at negative lags in occipital and thalamic regions and positive theta correlation with regions of the DMN (mostly the posterior cingulate and angular gyrus) near 0 lag. This latter finding mirrors our findings for alpha, and could be attributable to the fact that we utilized an eyes open resting paradigm, in contrast with Feige et al., who utilized an eyes closed paradigm. Gu et al. utilized an eyes-open paradigm at 2.1 sec TR and identified peak alpha-BOLD correlations near 0 sec lag for both posterior and anterior regions of the DMN (88% overlap). Their findings align with our findings in posterior regions of the DMN, although we find greater lags for anterior regions. This discrepancy could be attributable to differences in how alpha EEG was measured, since Gu et al. did not measure alpha relative to the aperiodic signal, and/or differences in study population, given our inclusion of participants with schizophrenia.

Methodological differences notwithstanding, these findings establish a body of evidence for noncanonical alpha-BOLD coupling within the DMN. However, underlying mechanisms remain unknown. Noncanonical coupling could relate to unexpected reversals of neurovascular coupling that are specific to the DMN, as seen during carbon dioxide challenge paradigms (13,14). This evidence is complemented by positron emission tomography (PET) studies that have revealed greater resting aerobic glycolysis (42), and dissociation between BOLD responses and cerebral metabolic rate in regions of the DMN (6). Overall, noncanonical neurometabolic coupling appears to be a defining characteristic of the DMN, and may prompt greater attention to energetic factors when interpreting DMN activity. As such, the DMN may represent an energetically favorable, stable state (43–45), in contrast with task-related frontal-parietal systems which may be more “costly” to engage (46). Free energy theories have proposed that the DMN operates to constrain endogenous spontaneous neural activity (47) which would demand less metabolic work overall (5). These perspectives are consistent with the everyday experience of mental effort (48): undirected free-association and daydreaming are “easy,” (akin to resting-state paradigms and typical participant instructions) whereas task-directed cognition is “hard” (47,49).

### Arousal and Stress

In addition to prior evidence for variability in EEG-BOLD coupling, our study was motivated by findings of increased latency of hemodynamic responses during stress in otherwise psychiatrically unaffected participants (15). Although our participants were not explicitly stressed via our resting-state paradigm, we found that the magnitude of the delay in alpha-BOLD coupling in the subgenual region of the ACC was correlated with self-reported measures of stress. Thus, we contribute confirmatory evidence for a hemodynamic lag related to stress, even in the absence of an experimentally induced stressor. Elbau et al. (15) interpreted their finding as being related to energy conservation and/or the need to reroute blood flow. Notably, their hemodynamic delays were largely associated with *negative* BOLD responses, suggesting a delay in the *reduction* of flow (i.e. unexpectedly sustained blood flow in the anterior DMN). Their interpretation assumes a passive neural-hemo relationship (hemodynamics as metabolic support for active neural activity), however, it is possible that their finding of sustained blood flow suggests an active, if not anticipatory mechanism, rather than a sluggishly evoked, canonical response, as we will discuss here.

Extending the hemo-neural hypothesis of Moore and Cao (18), we have argued that hemodynamic signals, in addition to providing metabolic support, can provide indirect (that is non-neurally mediated) information about ongoing behavior and arousal (16). Underlying mechanisms remain incompletely understood, however, this could occur via sensing of available metabolites in the form of ATP (50), hypoxia (51) and or mechanical transduction of blood pressure pulsations (52). Therefore, metabolic and hemodynamic signals may provide an alternative information pathway relevant to sustaining and/or maintaining a pattern of ongoing activity based on available energy. To distinguish this mechanism from canonical *e*voked BOLD signals in response to neural activity, we have described this process as a mechanism whereby BOLD activity *in*vokes ongoing neural activity. This hypothesis is supported by evidence that brainstem regions, particularly the locus coeruleus and other midbrain nuclei regulate cerebral blood flow as a mechanism to prepare for anticipated changes, such as stressors (53–56).

Notably, our non-hypothesized brain regions associated with anticipatory BOLD activity included a large cluster spanning the brainstem, midbrain and pons. Rather than seeing increased hemodynamic lags as a reaction to stress, here we propose that lags reflect an active, anticipatory mechanism, potentially linked to other slowly evolving neural signals (57,58) Hemodynamic lags are not only seen in stress paradigms, but also occur during the transition from sleep to an awake state. A recent study of sleep “inertia” (transient grogginess) demonstrated that thalamic BOLD lags alpha by ∼7.5 seconds, immediately after waking, and gradually returns to baseline of ∼2.5 seconds, 40 mins after awakening (59). Mirroring subjective mental effort under stress, BOLD lag during awakening could index requirements for greater mental work; waking up itself can at times, be reported as a stressful experience (60). Studies in non-human animals reveal similar dissociations between neural, vascular and metabolic responses, and particularly during transitions in arousal (7–10). Taken together, these findings point toward a fundamental association between subjective arousal and neurovascular coupling dynamics. More speculatively, rather than subjective arousal being exclusively encoded by specific neural networks and patterns of activity, we advance the idea that the felt sense of arousal, from grogginess to heightened stress, could be linked to the “tension” between neural activity and ongoing metabolic support, measurable by changing neurovascular coupling dynamics (49). Thermodynamically themed folk psychology metaphors such as “inertia” and everyday language around arousal (“moved,” “driven,” “dragging” (49)) are accurate in the sense that arousal transitions and stress states require metabolic energy to shift or maintain. This could also relate to work identifying ongoing neural activity as a dynamical system that must remain in motion (61,62). These hypotheses require study in human participants, along with closer investigation of the phenomenology of subjective mental energy.

### Neurovascular Coupling in Schizophrenia

Hemodynamic recruitment has long been identified as inefficient in SZP: executive networks are hyperactive during basic cognitive tasks, hypoactive during difficult tasks (63), and the default mode network is hyperactive (64). The hemodynamic response during task-based paradigms is delayed, particularly in the mPFC (19,20) and with a dysregulated release of dopamine in response to stress in these same regions (65). Our findings confirm neurovascular coupling abnormalities in medial prefrontal regions, by specifically measuring the lags between neural and hemodynamic correlations. These neuroimaging findings could be related to specific molecular and cellular deficits including neuroenergetic abnormalities (21), mitochondrial abnormalities (66), and mild ischemia in schizophrenia (67). Other potential mediators include potassium channel polymorphisms associated with deficient metabolism in the executive control network (ECN), delayed neurovascular coupling, and impaired neurovascular coupling during stress in animal models.(15,68,69) It is curious that stress, neurometabolic coupling delays and abnormalities in SZP all converge on the medial regions of the PFC (mPFC), including the sgACC, where the DMN and salience networks overlap (70). Delayed hemodynamic responses could relate to hyperactivity in the mPFC in SZ (71), that has been linked to aberrant salience attribution during self-referential processing (72). SZP could then interpret salient and stressful stimuli as threats, also explaining chronic, autonomic stress abnormalities in this population (73). Although our SZP and UAP did not differ in terms of reported stress, we also found no correlation between our coupling scores and heart rate, BMI or CPZ equivalents, all factors that are either direct mediators or moderators of the autonomic nervous system. Follow-up studies using stress paradigms, including direct assessment of the autonomic nervous system, and cardiovascular phenomena will be necessary to further explore this hypothesis. Lastly, it remains unknown if neurovascular coupling impairments in SZP reflect a chronic stress response or a more fundamental cellular-energetic deficit. In fact, these phenomena might be linked, given evolutionary evidence that increased cerebral metabolic demand and schizophrenia genetics co-evolved with systemic, metabolic changes early in hominid history (74–76).

### Implications for fMRI and Neuropsychiatric Conditions

fMRI is the lingua franca of neuropsychiatric brain research and typically relies on models of hemodynamic recruitment to parse differences in brain function between psychiatric populations. The presence of differences in neurovascular coupling in participants with SZP and that are linked to stress, should prompt more research to reconsider HRF as a potential confound in neuroimaging studies. For example, changes in resting-state functional connectivity could reflect regional changes in neurometabolic coupling. In part, this has been seen using so-called lag-analyses where the assumptions of near-instantaneous correlation are relaxed (77), with differences observed in adults with autism (78). Given the existence of new deconvolution methods, existing rsfMRI studies could be re-analyzed to examine the shape of HRF more explicitly. Impaired neurovascular function has also been linked to major depression (79) and depressive symptoms in brain trauma (80). More broadly, we may no longer be able to rely on the computational assumptions that molecular, cellular and network level metabolism is a passive support for neurons (81). Instead, metabolic factors are inextricably entangled within neural function. As such, neuroimaging methods, such as fMRI are highly relevant to neural function, but also provide a hemodynamic, “metabolic” window on neural function that should be seen as complementary to other methods that rely an electrical signals, such as EEG,

### Limitations and Future Directions

Despite strong pre-existing evidence for nonanonical neurovascular coupling in the DMN, this study was not designed to explicitly test this hypothesis, and our findings should be approached with caution. This study did not utilize newer, multiband sequences that could improve the temporal resolution of the BOLD signal. Further, we have not compared our findings to task based, event-related measures that are more commonly employed to estimate hemodynamic response latency. More generally, this study is limited by the anatomical resolution of fMRI and EEG. It is not a study of the cellular components of the neurovascular system, which includes neural, vascular endothelial and glial cells. Neural or metabolic changes, outside the resolution of the recording system may contribute to coupling that we are unable to observe (82). Along these lines, our brainstem findings are limited by the resolution of our 3T scanner. Precise anatomical designation necessitates high magnetic field strengths such as 7T which remains exploratory for simultaneous EEG-fMRI (83). Future studies might be complemented by arterial spin labeling, to derive greater specificity of arterial recruitment for neural activity dynamics in the fMRI scanner, as has been done in SZ (84). Our stress data was limited by the fact that not all participants took the stress questionnaire. In addition, this study was conducted at rest, so we were unable to test for stress effects explicitly. Future studies of stress and alongside autonomic measures of stress to determine the role of the ANS in this coupling phenomena.

## Data Availability

Data and code are available from the authors upon reasonable request.

## Acknowledgements

This work was supported by grants from the National Institute of Mental Health (R01MH058262–17 to JMF) and the VA (I01CX000497–06 and Senior Research Career Award to JMF, IK2CX002457 to MSJ). We gratefully acknowledge the input of our colleagues who provided feedback on an earlier draft of this manuscript: Parham Pourdavood, Kaia Sargent, and Terrence Deacon.

## Disclosures

Dr. Jacob reported grants from the VA during the conduct of the study. Dr. Mathalon reported grants from NIMH during the conduct of the study; consulting fees from Recognify Life Science, Syndesi Thera- peutics, Cadent Therapeutics, and Gilgamesh Pharmaceuticals, outside the submitted work. Dr. Ford reported grants from the VA and NIMH during the conduct of the study. No other disclosures were reported.

## Notes

### Competing Interest Statement

The authors have declared no competing interest.

### Funding Statement

This work was supported by grants from the National Institute of Mental Health (R01MH05826217 to JMF) and the VA (I01CX00049706 and Senior Research Career Award to JMF, IK2CX002457 to MSJ).

### Author Declarations

The IRB of the University of California, San Francisco and the San Francisco VA Healthcare System gave ethical approval for this work.

